# Microwave-Generated Steam Decontamination of N95 Respirators Utilizing Universally Accessible Materials

**DOI:** 10.1101/2020.04.22.20076117

**Authors:** Katelyn E. Zulauf, Alex B. Green, Alex N. Nguyen Ba, Tanush Jagdish, Dvir Reif, Robert Seeley, Alana Dale, James E Kirby

**Author notes:** Address correspondence to James E Kir.by. Katelyn E Zulauf and Alex B Green contributed equally to this work. Author order was determined based on seniority.

## Abstract

The SARS-CoV-2 pandemic has caused a severe, international shortage of N95 respirators, which are essential to protect healthcare providers from infection. Given the contemporary limitations of the supply chain, it is imperative to identify effective means of decontaminating, reusing, and thereby conserving N95 respirator stockpiles. To be effective, decontamination must result in sterilization of the N95 respirator without impairment of respirator filtration or user fit. Although numerous methods of N95 decontamination exist, none are universally accessible. In this work we describe a microwave-generated steam decontamination protocol for N95 respirators for use in healthcare systems of all sizes, geographies, and means. Using widely available glass containers, mesh from commercial produce bags, a rubber band, and a 1100W commercially available microwave, we constructed an effective, standardized, and reproducible means of decontaminating N95 respirators. Employing this methodology against MS2 phage, a highly conservative surrogate for SARS-CoV-2 contamination, we report an average 6-log_10_ plaque forming unit (PFU) (99.9999%) and a minimum 5-log_10_ PFU (99.999%) reduction after a single three-minute microwave treatment. Notably, quantified respirator fit and function were preserved, even after 20 sequential cycles of microwave steam decontamination. This method provides a valuable means of effective decontamination and reuse of N95 respirators by frontline providers facing urgent need.

**IMPORTANCE:** Due to the rapid spread of COVID-19 there is an increasing shortage of protective gear necessary to keep health care providers safe from infection. The CDC reports 9,282 cases of COVID-19 among U.S. healthcare workers to date (1). N95 respirators are recommended by the CDC as the ideal method of protection from COVID-19. Although N95 respirators are traditionally single-use, the shortages have necessitated the need for re-use. Effective methods of N95 decontamination that do not affect the fit or filtration ability of N95 respirators are essential. Numerous methods of N95 decontamination exist; however, none are universally accessible. In this study we describe an effective, standardized, and reproducible means of decontaminating N95 respirators using widely available materials. The N95 decontamination method described in this work will provide a valuable resource for hospitals, healthcare centers, and outpatients practices that are experiencing increasing shortages of N95 respirators due to the COVID-19 pandemic.

## INTRODUCTION

Since the initial cases in Wuhan, China in late December 2019, the COVID-19 pandemic, caused by the novel SARS-CoV-2 virus, has resulted in over 2.3 million infections and 160,000 deaths worldwide (2). Throughout this outbreak, the infection and resultant incapacitation of healthcare providers has been of significant concern. Each sick provider contributes to further nosocomial transmission and reduces the healthcare system’s capacity to handle incoming patient volume. One of the greatest threats to healthcare workers’ well-being is the critical shortage of personal protection equipment. Of particular concern are shortages in specialized N95 respirators.

The National Institute for Occupational Safety and Health (NIOSH) recommends N95 respirators for protection from particles <100nm in size, including viruses (3). Across the U.S. and worldwide, N95 prices have skyrocketed and supplies have dwindled. The Centers for Disease Control and Prevention (CDC) has released unprecedented guidance on the conservation, extended use, and limited re-use of N95 respirators in healthcare settings (https://www.cdc.gov/niosh/topics/hcwcontrols/recommendedguidanceextuse.html, https://www.cdc.gov/coronavirus/2019-ncov/hcp/ppe-strategy/decontamination-reuse-respirators.html). Effective, economical, accessible, and validated means of decontamination are urgently required.

Several N95 decontamination techniques have been validated and approved for clinical use. The CDC has summarized a variety of methods, including ultraviolet germicidal Irradiation (UVGI), ethylene oxide, vaporized hydrogen peroxide, moist heat incubation and microwave-generated steam (https://www.cdc.gov/coronavirus/2019-ncov/hcp/ppe-strategy/decontamination-reuse-respirators.html). The ideal method is one that is simultaneously rigorous enough to provide maximal decontamination and yet gentle enough to impart minimal structural damage to the N95 respirator. Microwave-generated steam is a uniquely promising method because of its potential for daily, affordable, and widespread use. Although microwave-generated steam has been shown to be effective in both decontamination and preservation of respirator function, the majority of published protocols rely on specialized commercial steam bags which are in limited supply today, or other unstandardized materials only available in research laboratories (4-8).

Here, as a quality assurance initiative at our institution, we set out to identify an N95 decontamination method that would allow repeated use of respirators. To assess decontamination, we utilized the *Escherichia coli* MS2 bacteriophage as a highly conservative surrogate for SARS-CoV-2. Here we describe development and evaluation of a simple, microwave steam decontamination protocol using affordable, readily available materials that achieves highly efficient disinfection of MS2 virus, while preserving respirator function for repeated re-use.

## RESULTS

To address the shortage of N95 respirators that are essential to keep healthcare workers protected from SARS-CoV-2, we set out to identify an effective method of N95 decontamination. Our goal was to find a decontamination method accessible to all practitioners in our distributed healthcare network, using MS2 phage as a model for SARS-CoV-2. Like SARS-CoV-2, the MS2 phage is a positive-sense, single-stranded RNA virus. Unlike SARS-CoV-2, the MS2 phage lacks a lipid envelope, making it more resistant to disinfection. Due to its stability, MS2 has been used to model disinfection of viruses such as norovirus and Ebola (9, 10).

Microwave-generated steam has proven to be an effective method of decontamination (4-8). In seeking a platform with widespread availability, we initially tested two protocols using common household items. Both protocols involved the use of a 10cm diameter ceramic mug filled with 60 mL water and covered with the mesh from a produce bag secured with a rubber band, on which the respirator was suspended directly above the generated steam (Figure 1A). In one assay, we placed the mug inside a ventilated gallon Ziploc bag. In the other assay, we place the mug directly in the microwave without containment. We then examined the ability of both methods to sterilize N95 coupons (excised 1 cm^2^ N95 fabric squares) inoculated with 10^7^ plaque forming units (PFU) of MS2 phage. The inoculation of 10^7^ PFU represents a higher viral load than any viral droplet a healthcare provider is likely to encounter in the clinical setting (11). After 1 minute of microwave steam decontamination, we saw no significant difference in MS2 phage reduction between the two methods (Figure 1B). It is important to note that both methods resulted in greater than a 4-log_10_ reduction in MS2 titer after only 1 minute of microwave treatment. The Ziploc bag, however, melted under this treatment and posed the risk of steam burns during retrieval of the N95 respirator. Noting equal efficacy, we proceeded with the open container method of steam decontamination for all further work.

**Figure 1:**
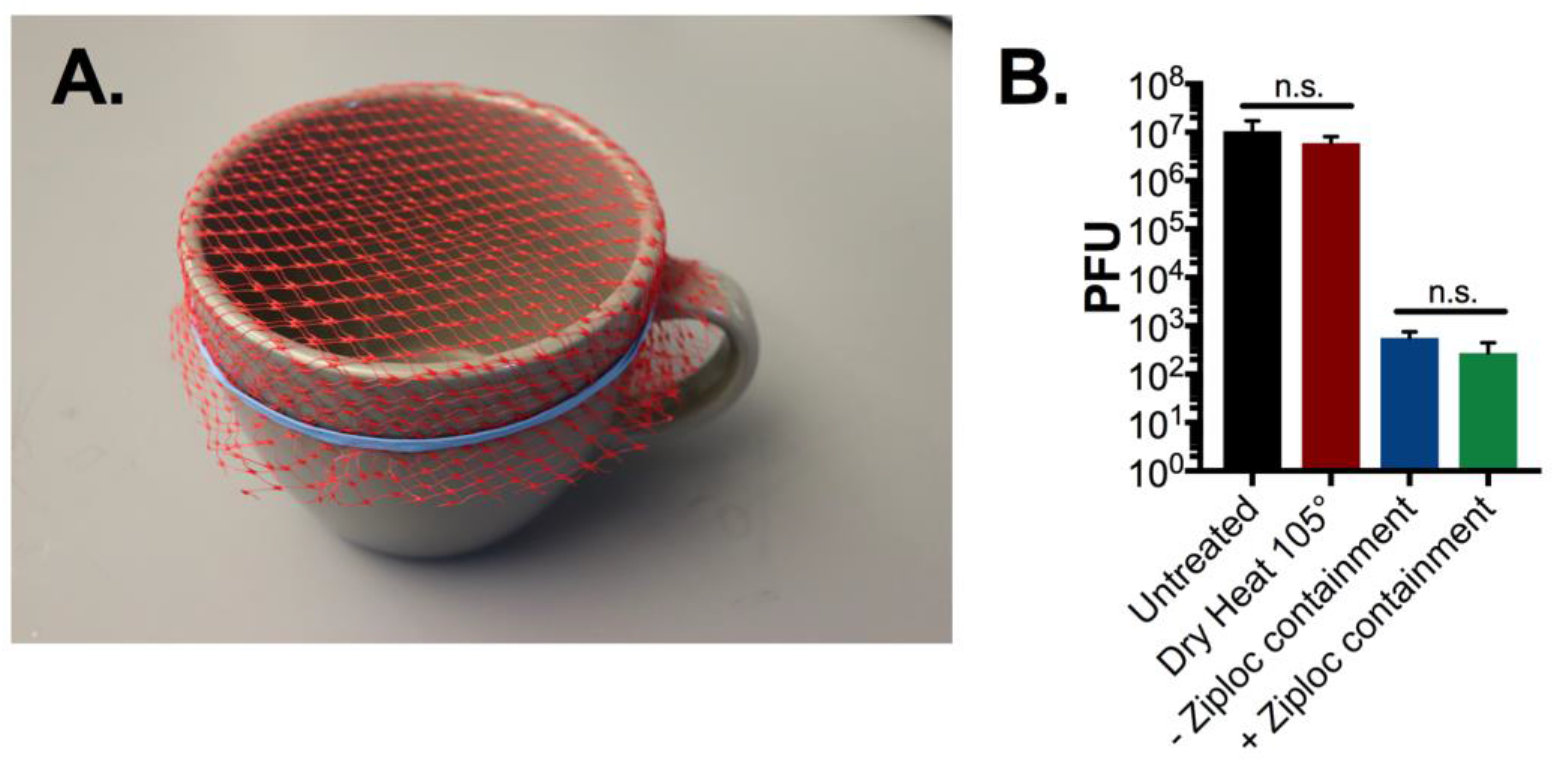
N95 microwave steam decontamination by ceramic mug either inside or in the absence of Ziploc containment. **(A)** Image of ceramic mug decontamination system. A 10 cm diameter mug was filled with 60 ml of water and covered with mesh from a produce bag, secured with a rubber band. Triplicate N95 1cm^2^ coupons were placed on top of the mesh. The mug was then placed in the microwave either in a sealed, ventilated Ziploc bag or directly into the microwave. **(B)** After a 1-minute microwave treatment, with or without Ziplog bag enclosure, or a 60-minute treatment with dry 105°C heat, phage was extracted from N95 coupons and quantified by plaque assay. Triplicate untreated N95 coupons were included as controls in all assays. There was no significant reduction in plaque titer between Ziploc-enclosed and open mug decontamination systems or between dry-heat treated and untreated controls (*p* = 0.9 or *p*=0.66 respectively as determined by ANOVA with Holm Sidak post-hoc test). PFU = plaque forming units, a direct measure of viable viral titer.

To identify the optimal length of microwave time required for MS2 phage decontamination, we performed a dose-response test using 1 minute increments where we examined the decontamination of 10^7^ PFU of MS2 on 1cm^2^ N95 coupons placed over an open mug (as described above). Following three minutes of microwave steam treatment there were no detectable MS2 phage remaining on the coupons (Figure 2A). To accurately assess the ability of this method to decontaminate all areas of an N95, we inoculated 10^7^ PFU of MS2 phage onto ten discrete sections of an N95 respirator (Figure 2C). Following three minutes of treatment, we observed a greater than 4-log_10_ reduction in PFU on all N95 respirator segments, except the elastic straps which only showed a 1-3 log_10_ reduction in PFU (Figure 2D). Due to the limited diameter of the mug, the elastic straps draped over the edges, and presumably were minimally exposed to microwave-generated steam (Figure 2B). Consequently, we hypothesized that direct exposure to steam is essential for effective decontamination, and sought to identify a commercial container of sufficient diameter to treat an entire respirator.

**Figure 2:**
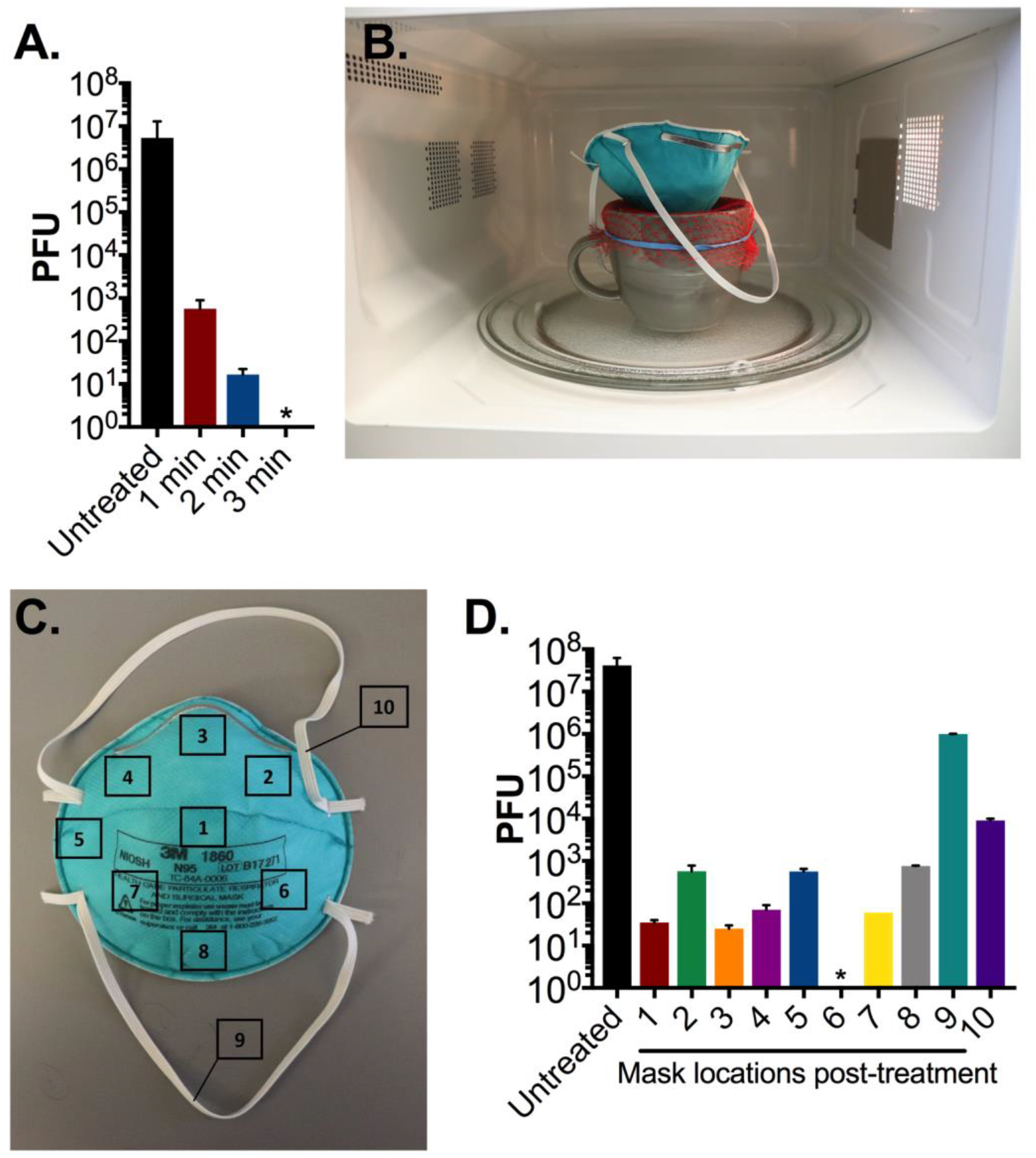
N95 decontamination by microwave generated steam over an open ceramic mug. **(A)** Triplicate N95 coupons treated with 10^7^ PFU MS2 were placed on the mesh covered ceramic mug and treated for the indicated durations in an 1100W microwave. After treatment, phage was extracted from N95 coupons and quantified by plaque assay. **(B)** We next evaluated treatment of an entire N95 respirator on the mug decontamination system. **(C)** 10^7^ PFU of MS2 was spotted on 10 pre-marked sections of a whole N95 respirator as indicated. **(D)** After a 3-minute treatment in an 1100W microwave demarcated pre-treated segments measuring 1 cm^2^ were excised from the respirator, and MS2 phage was then extracted and quantified by plaque assay. Triplicate untreated pre-cut N95 coupons were included as a control in all assays. Bars shown are mean and standard deviation of phage titers from each excised segment from a single respirator. * indicates no viable MS2 detected. Limit of detection of all assays is 10 PFU. Data shown are representative of three separate respirator experiments.

Ultimately we selected a generic glass container sized at 17 × 17 × 7.5 cm (LxWxH) that had an opening large enough to expose the entire N95 respirator to the vertical column of generated steam. As with the ceramic mug, we secured mesh from a produce bag over the top of the container with a rubber band and added 60ml of water to the basin (Figure 3A-B). We repeated a sterilization time course against 1cm^2^ N95 respirator coupons in 1-minute increments. After 2 minutes of microwave steam treatment we were unable to detect residual viable phage on the coupons (Figure 3C). This represents a 1 minute reduction in sterilization time compared to the ceramic mug decontamination assay, indicating that the glass container is a more efficient decontamination system.

**Figure 3:**
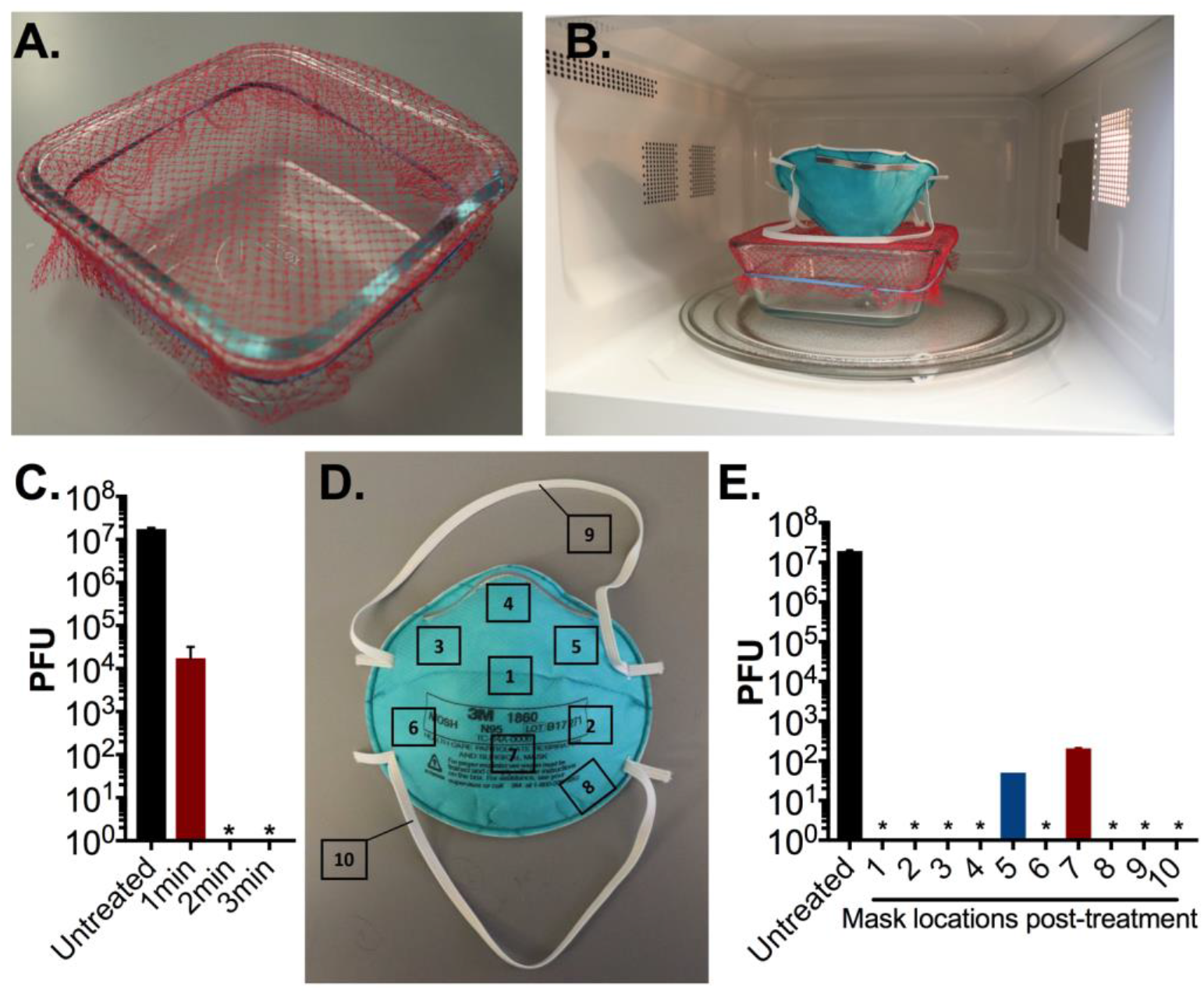
N95 decontamination with microwave-generated steam over an open glass container. **(A-B)** Image of glass container decontamination system. A 17 cm × 17 cm glass container was filled with 60 ml of water, covered with mesh from a produce bag, secured with a rubber band. **(C)** Triplicate N95 respirator coupons inoculated with 10^7^ PFU MS2 phage, placed on the mesh-covered container, and treated for indicated times in an 1100W microwave. After treatment, MS2 phage was extracted from N95 coupons and quantified by plaque assay. **(D)** 10^7^ PFU of MS2 phage was spotted on 10 different pre-marked locations on a N95 respirator as indicated. **(E)** The whole N95 respirator was then treated for 3 minutes as shown in Fig. 3B in an 1100W microwave. Demarcated segments measuring 1 cm^2^ encompassing the area of inoculation were excised from respirator, and MS2 phage was extracted and quantified by plaque assay. Triplicate untreated pre-cut N95 coupons were included as a control in all assays. Data shown are the mean and standard deviation of plaque titers from a single respirator and are representative of three separate experiments. In one experiment, no viable PFU were detected from all excised segments (data not shown). * indicates no viable MS2 was detected. Limit of detection of all assays is 10 PFU.

We next examined the ability of the glass container to sterilize a whole N95 respirator. As described above, we inoculated ten sections of an N95 respirator with MS2 phage and treated the respirator for 3 minutes over the container system (Figure 3D). Only 20% of the sampled sections exhibited residual phage, and of those, each exhibited a 5-log_10_ reduction in viable phage (Figure 3 E). In one of three trials on separate N95 respirators, there was complete sterilization on all sampled sections with no detectable PFU remaining post-treatment. Importantly across all assays, viral load was reduced by an average of 6-log_10_ PFU (99.9999% with a minimum of 5-log_10_ PFU (99.999%) reduction. These results indicate that open container treatment is an effective method of N95 decontamination.

Since it is essential that any decontamination method not reduce the filtration or integrity of N95 respirators, we examined N95 respirator fit and function after sequential treatments. After 1, 5 or 20 three-minute treatment cycles, no damage was evident in the integrity of the respirators or their component parts (i.e. straps, foam fittings, nosepiece). Additionally, no odors were detectable post-treatment, which is consistent with previous reports of microwave generated steam decontamination (5). Furthermore, Quantitative Respirator Fit Testing conducted with a Portacount Fit Tester 8030, in keeping with the Occupational Safety and Health Administration’s (OSHA) procedures (12), did not demonstrate any changes in respirator performance after 1, 5, or 20 treatment cycles. Across all seven exercises in the OSHA-accepted fit test protocol, we observed component Fit Factor scores in excess of the 100 Fit Factor minimum and overall Fit Factor values >175 after 1, 5, and 20 treatment cycles. Cumulatively these data indicate that fit, seal, and filtration of the N95 respirator is preserved even after 20 consecutive treatments.

Lastly, post-treatment respirators did not show a significant change in mass (< 1mg) compared with pre-treatment respirators. Therefore, in contrast to another study employing commercial microwave steam bags, microwave-generated steam over an open vessel was not associated with significant water retention (6). This is likely due to the fact that the N95 respirator is suspended above the steam and is not in any sustained, direct contact with water. The lack of water retention means little to no drying time is required post-treatment prior to N95 respirator use.

In summary, we identified an effective method of N95 decontamination by microwave generated steam utilizing universally accessible materials. Our method resulted in almost complete sterilization after only three minutes of treatment and did not appear to affect the integrity of N95 filtration or fit with repeated treatment.

## DISCUSSION

Due to the rapid spread of COVID-19, hospitals, healthcare centers, and outpatient practices are experiencing increasing shortages of protective gear necessary to keep health care providers safe from infection. Specialized N95 respirators are recommended by the CDC for protection from COVID-19 (https://www.cdc.gov/niosh/topics/hcwcontrols/recommendedguidanceextuse.html.https://www.cdc.gov/coronavirus/2019-ncov/hcp/ppe-strategy/decontamination-reuse-respirators.html). Although N95 respirators are normally only recommended for single use, the severe shortages have necessitated the need consistent for re-use. During patient care, however, the surfaces of N95 respirators are likely contaminated by viral aerosols. Recent work has demonstrated that SARS-CoV-2 can survive on surfaces for up to 72 hours (13). Without decontamination, N95 respirators can serve as infectious fomites and pose a risk to healthcare providers. In order to conserve supply and provide healthcare workers protection, there has been an effort to identify viable methods of N95 decontamination.

The CDC reports a variety of decontamination methods, including ultraviolet germicidal Irradiation (UVGI), ethylene oxide, vaporized hydrogen peroxide, moist heat incubation and microwave-generated steam https://www.cdc.gov/coronavirus/2019-ncov/hcp/ppe-strategy/decontamination-reuse-respirators.html). However these methods have significant limitations. UVGI is limited by inherent shadow effects of a light-source, and variability in dosages due to bulb age and differing platform constructions (14). Ethylene oxide is efficacious in eradicating microbial contamination, but is also a known carcinogen and teratogen, and exposure has been correlated with neurologic dysfunction (15). Vaporized hydrogen peroxide (“VHP”) is highly effective, killing greater than 99.9999% of surrogate microbial contaminants (a 6-log_10_ reduction), while preserving respirator filtration function (16). Yet the technology necessary for VHP is limited to larger healthcare systems that can afford the required equipment. Therefore, there is an urgent need for viable methods of decontamination that are safe, effective, and available in diverse clinical settings.

In order to identify a generally accessible N95 respirator decontamination method, we focused our efforts on microwave generated steam decontamination. Microwaves are ubiquitous and previous studies have demonstrated the effectiveness of microwave steam decontamination. To-date, however, studies examining microwave generated steam decontamination have employed both specific and laboratory-generated materials (e.g. pipette-tip boxes, modified reservoirs, commercial steam bags, etc.) that may not be generally available or easily reproduced (4-8).

The goal of this work was to identify a widely accessible, microwave-generated steam decontamination method. To this effect, we only utilized common household items. We first examined whether contained steam was a more effective than an open steam vessel. As commercial microwave sterilization bags have previously demonstrated efficiency, we examined if common Ziploc bags might provide similar benefits (6). Although Ziploc-enclosed decontamination was effective, our results indicated that it is a more cumbersome system and containment could be dispensed with entirely. Furthermore, Ziploc bags began to melt when exposed to more than one minute of microwave generated steam and posed the risk of thermal burns from contained steam making the enclosed method less compelling.

With additional study, we found that use of a generic glass container measuring 17×17×7.5cm (LxWxH) resulted in the most efficient and practical N95 respirator decontamination system. Using this method, we observed almost complete sterilization of the N95 respirator after a single 3-minute treatment. On average, we found a 6-log_10_ reduction in viable MS2 phage with a minimum of a 5-log_10_ reduction. During decontamination treatments, we positioned the N95 respirator with its convex surface pointed downward, onto the mesh-covered container, maximizing steam exposure. Placement was otherwise made without regard to specific orientation of the respirator, simulating real world application. Post-treatment water retention by the N95 was undetectable, eliminating a need for drying time before reuse. Importantly, this method was validated for use of 20 times on a single respirator without detrimental effect on respirator integrity or fit. In contrast, a recent preprint demonstrated that fit and seal integrity was compromised in UV- and heat-treated N95 respirators after 3 treatment cycles, and in ethanol-treated respirators after 2 treatment cycles (17). Given these findings, decontamination by microwave-generated steam may provide an ideal solution for broad N95 respirator re-use, with minimal treatment duration, minimal post-treatment processing, and maximal re-use potential.

The MS2 bacteriophage was used as a model of SARS-CoV-2 in this study. It provided a facile system for rapid quantitative evaluation of respirator disinfection. Importantly, MS2, similar to SARS-CoV-2, is a positive-sense single stranded RNA virus. However there are some important distinctions. MS2 is a non-enveloped virus encased in a protective icosahedral protein capsid. SARS-CoV-2, in contrast, is a lipid enveloped virus, making it more susceptible to disinfection methods. Previously, MS2 has been used as a surrogate for protein capsid-protected noroviruses and also for enveloped Ebola virus, and has been reported to be significantly more resistant to disinfectant than both (9, 18). These differences were highlighted in our present study by observation of complete resistance of MS2 to dry heat inactivation at 105°C for 60 minutes (Figure 1B) compared with previously reported complete inactivation of SARS-CoV-2 by 70°C dry heat treatment for the same duration (17). Therefore, the substantial reduction in viable MS2 by microwave steam decontamination gives confidence in an appropriate safety margin for SARS-CoV-2 decontamination and suggests the additional benefit of disinfection of other viruses that may also contaminate respirators during re-use.

Our N95 decontamination system uses only commonly available materials: a glass container, mesh from a produce bag that can be found at any grocery store, and a rubber band, as well as a common household 1100W or 1150W microwave. All microwaves used in this study had a turntable to enable rotation while heating (Figure 1B-C, 3A), a feature that likely promotes uniform heating and steam production. It is important to note that the microwave treatment did not result in sparks even when there was metal present on the N95, which is consistent with previous reports (5). Our decontamination protocol was validated with a glass container, and it is possible that steam generation and results would be affected using containers made of other materials.

We began our study using a ceramic coffee mug to generate steam. However, there were several limitations to this method. Notably, sizes of ceramic mugs vary widely, making this method hard to standardize. Although we observed complete sterilization of the 1 cm^2^ coupons on the ceramic coffee mug, we did not observe a similar result using the entire N95 respirator (Figure 2) where significant quantities of viable virus remained post-treatment (10^2^-10^6^ PFU). In contrast, the larger surface area provided by the glass container led to almost complete sterilization at all locations. Furthermore, the 10 cm diameter mug opening was still too small to suspend the elastic straps of the respiratory above the mug orifice. This led to inefficient decontamination of the elastic straps, in contrast to findings using the larger glass container. Our study highlights the need to examine the whole respirator for disinfection rather than just small coupons isolated from the filtration material as has been generally performed in N95 respirator disinfection studies.

Taken together, this work demonstrates the effectiveness of an affordable, simple method of N95 respirator decontamination. Use of common household items and the ability to re-sterilize the respirator at least 20 times without detriment to filtration or fit provides a compelling disinfection method that should prove generally accessible to diverse settings including outpatient practices, frontline providers, and remote clinical settings.

## MATERIALS AND METHODS

### Generation of High Titer MS2 Phage Lysate

MS2 bacteriophage were recovered from cultures of *Escherichia coli* strain W1485 using standard phage isolation techniques (19). We added 2 mL of 10^9^ PFU/mL MS2 to a 50 mL culture of *E. coli* W1485 in exponential phase (OD ∼ 0.2). Overnight growth resulted in MS2-mediated lysis of growing cells. This suspension was centrifuged at 4000 g for 10 minutes and filtered through a 0.22 micron polyethylene filter. The filtrate was then assessed for viral titer and adjusted to a final concentration of 10^9^ PFU/mL for all downstream experiments.

### N95 Respirator Decontamination

Decontamination tests were conducted using 3M N95s (Model 1860 Health Care Particulate Respirator and Surgical Mask). Excised 1cm^2^ coupons or whole N95 respirators were treated with 10 µL of 10^9^ PFU/mL MS2 resulting in a ∼10^7^ PFU inoculation. When a whole respirator was treated, ten sections were demarcated and inoculated, including two spots on the elastic straps. The inoculated N95 coupons and whole respirators were then allowed to dry inside a biosafety cabinet for 2 hours. Once dried, pre-cut triplicate N95 respirator coupons were removed to quantify viral load prior to intervention.

Mesh from produce bags (multiple variants were utilized in this study) were secured across the top of either a ceramic mug or glass container with rubber bands. The mug utilized in this study was 10cm in diameter. The glass container (Snapware 4-cup Food Storage Container made with Pyrex Glass) is roughly 17cm x17cm x7.5cm (6.5×6.5×3in). Both the mug and glass container were filled with 60ml (¼ cup) of water for steam generation. N95 coupons and respirators were placed outward facing side down, onto the mesh, for direct suspension above the steam (Figure 1C, 3B). When a Ziploc bag was utilized, a 2 cm slit was cut in the upper-right side of the bag to vent excess steam. The N95 respirator coupons and whole respirators were then treated for the indicated time in either a 1150W or a 1100W microwave. Both microwaves utilized in this study contain a turntable for uniform heating. It is noteworthy that results were consistent between both microwaves used in this study.

For dry heat treatment, 1cm^2^ N95 respirator coupons were exposed to 105°C dry heat for 60 minutes in a hybridization oven.

### MS2 phage quantification

Viral load on pre- and post-intervention N95 respirators and coupons was measured using established plaque assay protocols (19, 20). For whole respirators, each treated area was cut and removed from the N95 and processed individually. Each piece was submerged in 1 mL LB broth and vortexed for one minute to elute MS2 phage. The LB suspension was then serially diluted, following which 100 µL from each of the phage dilutions was mixed with 100 µL of exponential phase host *E. coli* W1485 cells. This mixture was then vortexed with 3 mL of LB top agar (0.6% agar w/v) and spread on LB agar plates (1.5% agar w/v). All plates were incubated at 37°C overnight to allow for bacterial lawns and viral plaques to grow. Plaques were quantified and the total PFU burden was calculated. The limit of detection of this assay was 10 PFU.

### N95 Respirator Filtration and Fit testing

N95 respirators treated 1, 5 and 20 consecutive times were examined for overall integrity and filtration performance. There were not any observed changes in comfort, breathing effort, or odor between control and microwave-treated respirators. Quantitative respirator fit testing was conducted using a PortaCount Pro 8030 Fit Tester (TSI incorporated). Testing was conducted in accordance with the definitions, thresholds, and protocols for respiratory protective equipment outlined in OSHA 29CFR1910.134 (https://www.osha.gov/laws-regs/regulations/standardnumber/1910/1910.134). Sodium chloride was used as a non-hazardous test aerosol using a TSI particle generator. Fit factor was calculated automatically on the PortaCount Pro 8030 Fit Test. Fit Factor represents the ratio of the average ambient aerosol concentration to that measured inside the respirator during each exercise. For testing, a sample port was installed in the breathing zone of the respirator using N95 fit test adaptors. The respirator was donned for five minutes before the quantitative test to check for adequacy of respirator fit, perform user seal checks, and purge particles trapped inside the respirator. The fit-test entailed the seven OSHA-approved exercises, each up to one minute in duration, including: Normal Breathing, Deep Breathing, Head Side to Side, Head Up and Down, Talking, Grimacing (15 seconds only), and Bending Over. The PortaCount Pro Fit Tester calculated the Fit Factor for each exercise, as well as an overall, averaged Fit Factor. Passing entailed a minimum Fit Factor of 100 on individual exercises and overall score alike. In keeping with the operational manual, the PortaCount Pro Fit Tester underwent daily maintenance checks throughout the period of testing, to ensure continuous quality monitoring.

### Water Retention Studies

We quantified water absorption into N95 respirators during the microwave steam decontamination treatment as described previously (6). We measured the mass of N95 respirators pre-treatment and immediately post-treatment using an analytical balance. We subtracted the pretreatment mass from the post-treatment to quantify the amount of water absorption. In all assays, there was <1mg difference between the two measurements, which remained constant even with multiple treatment cycles.

## Data Availability

All data are provided within the manuscript.

## ACKNOWLEDGEMENTS

We would like to thank the laboratory of Michael Baym at Harvard Medical School for the generous gift of the MS2 phage and the host *E. coli* strain and Thea Brennan-Krohn for critical review of the manuscript. KEZ was supported by the National Institute of Allergy and Infectious Diseases training grant (T32AI007061). The content is solely the responsibility of the authors and does not necessarily represent the official views of the National Institutes of Health.

## Notes

### Competing Interest Statement

The authors have declared no competing interest.

